# Derivation of anthropometric-based equations to predict lean body mass composition of cancer patients

**DOI:** 10.1101/2021.07.05.21259938

**Authors:** Autumn B. Carey, Ashley S. Felix, Jared D. Huling, James B. Odei, Christopher C. Coss, Xia Ning, Macarius M. Donneyong

## Abstract

**Background:** Lean body mass (LBM) composition of cancer patients is a predictor of chemotherapy-related adverse events and overall cancer survival. However, clinicians lack validated algorithms that can be applied to measure the LBM of cancer patients to facilitate accurate chemotherapy dosing. Our goal was to develop LBM predictive equations using routinely measured anthropometric measures among cancer patients.

**Methods:** We leveraged the 1999-2006 National Health and Nutrition Examination Survey (NHANES) data cycles containing information on self-reported cancer diagnosis, LBM measures based on dual-energy x-ray absorptiometry (DXA) and several anthropometric and demographic factors. We restricted our analysis to participants who had been diagnosed with cancer at the time of surveys. The data was randomly split to 75%:25% to train and test predictive models. Least absolute shrinkage and selection operator (LASSO) models were used to predict LBM based on anthropometric and demographic factors, overall and separately among sex and sex-by-race/ethnic subgroups. LBM measured directly with DXA served as the gold standard for assessing the predictive abilities (correlations [R^2^] and the Root Mean Square Error [RMSE]) of the derived LBM-algorithms. We further compared the correlations between both DXA-based LBM and predicted LBM and urine creatinine levels, a known biomarker of muscle mass.

**Results:** We identified 1,777 cancer patients with a median age of 71 (interquartile range [IQR]: 60-80) years. The most parsimonious model comprised of height and weight, which accurately predicted LBM overall (R^2^=0.86, RMSE =2.26). The predictive abilities of these models varied across sex-by-race/ethnic groups. The magnitude of correlations between derived LBM-algorithm and urine creatinine levels were larger compared to those measured between DXA-based LBM and urine creatinine levels (R^2^=0.30 vs. R^2^=0.17)

**Conclusions:** We successfully developed a simple sex-specific and sex-by-race/ethnicity-specific models to accurately predict the LBM of cancer patients by using only height and weight. The simplicity and high accuracy of these models make them inexpensive alternatives to measuring the LBM of cancer patients. Data on the LBM of cancer patients could help guide optimal chemotherapy dose selection among cancer patients.

## INTRODUCTION

In 2020, approximately 1.8 million cancers are expected to be diagnosed in the United States, with approximately 0.6 million cancer-related deaths^1^. Cancer is the second leading cause of death with a five-year mortality rate of 34.1%^1, 2^. Notwithstanding this high mortality burden, advances in treatment underlie significant declines in cancer mortality over the past 20 years, with rates dropping from 209.9 per 100,000 people in 1995 to 158.7 per 100,000 people in 2015^2^. Chemotherapy has been shown to improve cancer survival, especially for advanced stage cancers^3, 4^. However, this benefit could be offset by the negative impact of chemotherapy-induced adverse events and chemotoxicity effects, such as hair loss, nausea, vomiting, or death. These adverse effects can lead to treatment cycle interruption or discontinuation, both of which contribute to poor cancer outcomes and higher mortality^5^.

Chemotoxicity is caused by the excretion or poor metabolism of chemotherapeutic drugs, resulting in higher than expected circulating levels of the drug ^6, 7^. Known risk factors for poor chemotherapeutic drug metabolism are determinded by the pharmacokinetic properties of the chemotherapeutic drug, but often include low lean muscle mass^6^. Prado et al (2007) found that a cut point of 20mg 5-fluorouracil per kilogram of total lean body mass (LBM) is a threshold for developing toxicity (p-value<0.01)^6^. LBM is comprised of metabolic tissue (liver and kidneys), skeletal muscle, intracellular water, extracellular water, and bone^8^. Thus, research continues to emphasize the value of measuring body composition to normalize drug dosing, optimize outcomes, and mitigate chemotoxicty ^6, 9-12^.

In clinical practice, chemotherapy dosing is based on body weight, body mass index (BMI) or body surface area (BSA). This practice may pose challenges to measuring the optimal dose of chemotherapy for individual patients. BMI is limited as it does not capture a valid measure of body composition phenotypes (i.e. lean muscle mass, lean muscle mass percentage, fat mass, and fat mass percentage) and does not readily explain inter-patient variability in drug clearance inter-patient variability despite its use in many epidemiological studies assessing chemotoxicity risk and cancer outcomes^13,14^. Chemotherapy dosing based on BSA is a common clinical practice in oncologic settings. However, variation in the pharmacokinetics of most chemotherapies has failed to be standardized by BSA-based dosing^15-17^.

Dual-energy x-ray absorptiometry (DXA) and computed tomography (CT) are imaging tools used to accurately calculate body composition among cancer patients. However, CT scans are expensive and emit a high dose of ionizing radiation^18^. DXA is relatively inexpensive and emits low amounts of radiation, but whole body DXA examination equipment is not typically accessible in an oncologic clinic setting^18, 19^. Due to these limitations, anthropometric measures in conjunction with age, sex, weight, and height are needed to precisely estimate body composition for chemotoxicity risk stratification ^13, 20^.

Several studies have shown that LBM can be estimated accurately by using equations that comprise of simple anthropometric measures ^21-24^. However, none of these studies published data the application of anthropometric-based equations to measure LBM specifically among cancer patients. Therefore, our primary objective was to develop anthropometric-based models to accurately predict LBM among cancer patients.

## METHODS

### Study Population

Participants were selected from the National Health and Nutrition Survey (NHANES), a series of cross-sectional surveys designed to assess the health and nutritional status of adults and children in the United States conducted by the Centers for Disease Control and Prevention^25^. Because NHANES is a publically available dataset, The Ohio State University IRB did not constitute the project to require human subjects review. NHANES participants were selected using complex, stratified, multi-staged probability sampling to be representative of the US civilian non-institutionalized population^26-28^. All survey cycles between 1999 and 2014 were utilized. **Figure 1** depicts sample selection for the development of the final training, testing and validation of models.

**Figure 1.**
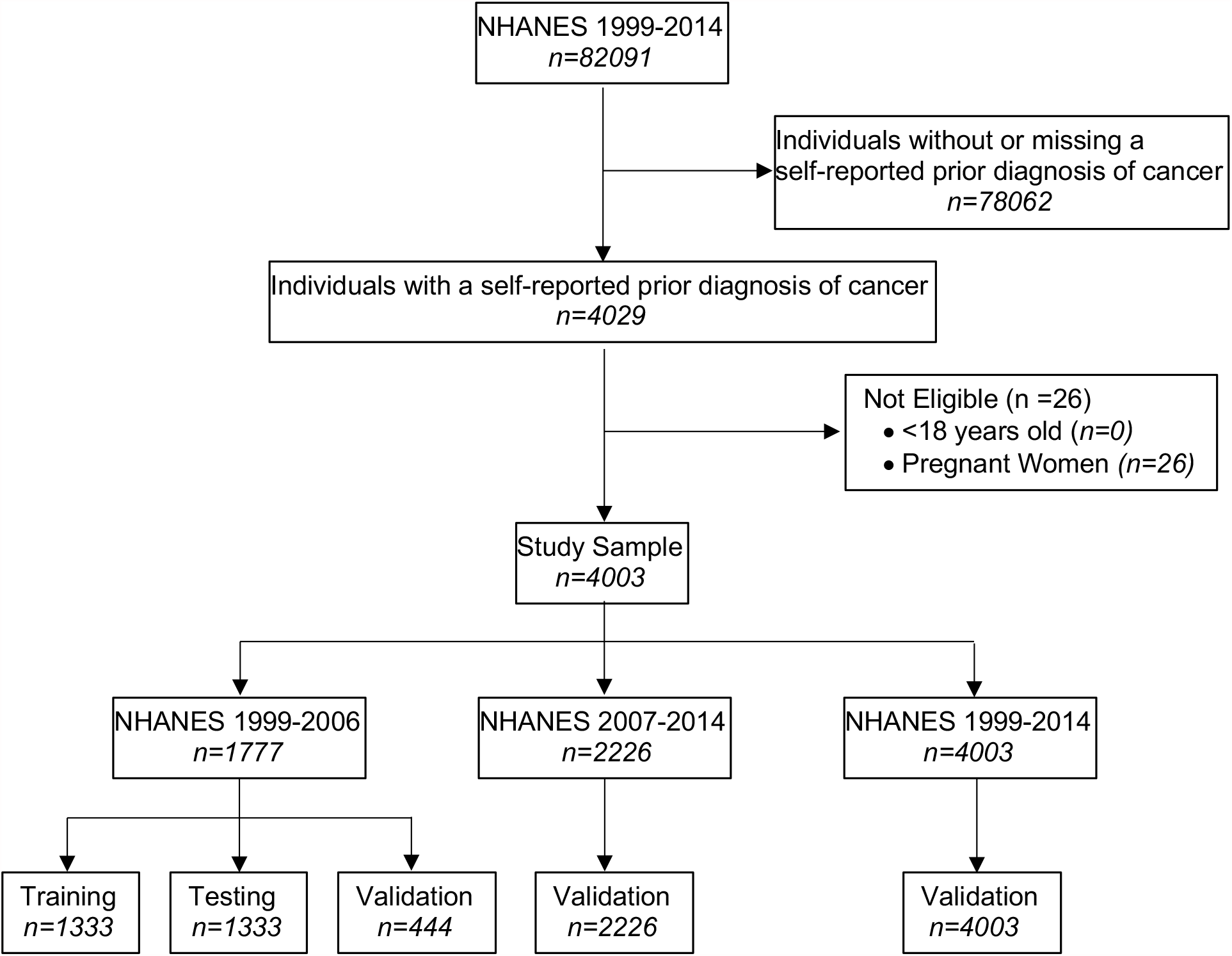
Flow chart describing the development, training, and validation samples for analysis from 8 NHANES cycles 1999-2014

**Figure 3.**
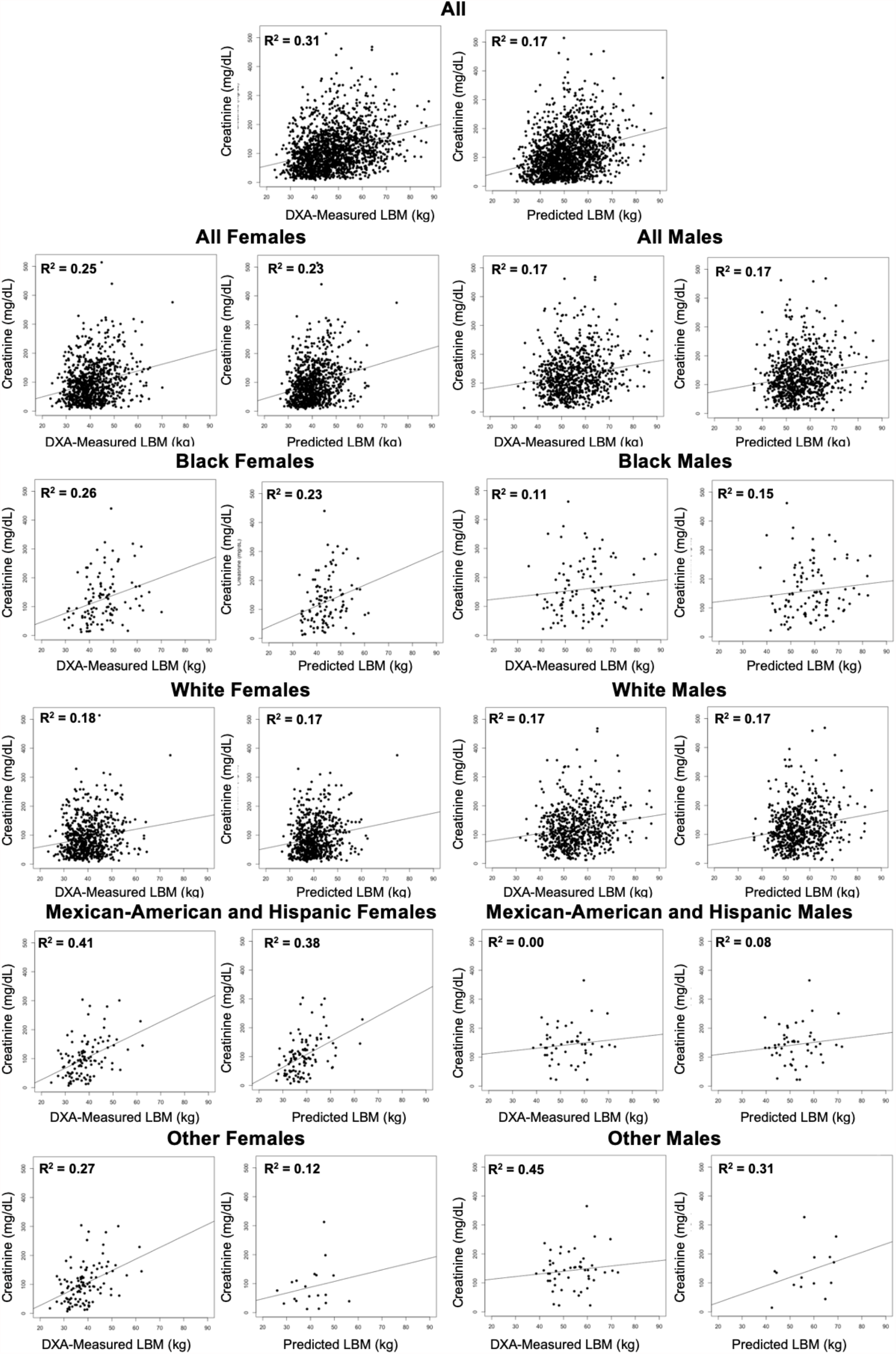
Correlations between serum creatinine levels (mg/dL) with DXA-based LBM (kg) and derived LBM-algorithm (kg) in the overall sample and by sex and race/ethnicity subgroups.

#### Inclusion/Exclusion criteria

We included participants with a self-reported cancer diagnosis aged ≥18 years old. Pregnant women were excluded^29^.

### Outcomes

#### Dual-Energy X-Ray Absorptiometry Measurements

Whole body DXA scans were used to measure LBM, fat mass, and fat mass percent of NHANES participants. A Hologic QDR 4500A fan-beam densitometer within the NHANES Mobile Examination Centers was used to measure body composition^30-32^. Details of the body composition measurement and exclusion of participants have been previously published^31^. Outliers at the individual level were coded as missing by NHANES staff prior to completing statistical analysis^32^.

### Predictors

#### Anthropometric Measurements

We focused on the following anthropometric measures that have been assessed as predictors of LBM in the published data ^21-24^: height (cm), weight (kg), arm circumference (cm), abdominal circumference (cm), calf circumference (cm), thigh circumference (cm), tricep skinfold (cm), and subscapular skinfold (cm). These anthropometric measures were collected in the Mobile Examination Center by two trained health technicians, an examiner and recorder, per NHANES standardized procedures and equipment^33^. For the modeling procedures, all anthropometric variables were modeled as continuous variables, and tricep and subscapular skinfold measures were converted from units of millimeters to centimeters.

#### Other measures

Demographic factors examined in this study are age, time between cancer diagnosis and NHANES screening age (years), race (Black Non-Hispanic, Mexican American and Hispanic, White Non-Hispanic and other (which includes multiracial), and sex (male and female). Age was top coded at 80 years old in NHANES cycles 1999-2006 and 85 years old in NHANES cycles 2007-2014 to maintain confidentiality. Thus, for this analysis, all individuals over the age of 80 were top coded at 80 to be uniform across all survey cycles. Additionally, age of cancer diagnosis or most recent cancer diagnosis,if multiple diagnoses were reported, were examined to calculate the time between cancer diagnoses and LBM measurement. In addition to these demographic factors, we selected urine creatinine levels as a potential biomarker for muscle ^34, 35^. Urine creatinine levels were measured via 24-hour urine samples collected within 2 weeks of DXA measurements and analyzed by NHANES staff ^36^.

### Statistical Analysis

The complex sampling design of NHANES was accounted for by proper use of the sample and cycle weights in all analysis^37^. We implemented the analysis described below with SAS 9.4 and R 1.2.1335.

#### Descriptive analysis

Proportions and means were used to describe categorical and continuous variables, respectively, overall, by sex, and by NHANES data cycles (with and without whole-body DXA measurements).

#### Missing Data

Greater than 10% of participants included in our analytic sample were missing data for BMI (kg/m^2^), BSA (m^2^), all eight anthropometric measures, creatinine (mg/dL), and DXA measures. We applied Multivariate Imputation by Chained Equations (MICE) models^38^ to impute all missing values of predictors. MICE requires missing data models to perform the imputation, for which flexible random forest models were used due to their favorable performance^39^. To obtain valid estimates that take into account the uncertainty in imputation, we construced twenty-five imputed datasets were constructed. NHANES had already imputed missing DXA values prior to making data available to researchers by using multiple-imputation methodology by the National Center for Health Statistics (NCHS)^32^. This missing data imputation was based on a sequential regression multivariate imputation methodology to reflect smaller standard errors, narrower confidence intervals, and falsely failing to reject null hypotheses that are often reflected when utilizing single imputation methodsDXA technical doc^32^.

#### Algorithm development

We implemented five different models based on a combination of anthropometric and demographic factors among the overall sample: 1) Model 1: height (cm) and weight (kg) only; 2) Model 2: Model 1 plus age, time between cancer diagnosis and LBM measurement date, sex, race/ethnicity; 3) Model 3: Model 2 plus interaction terms between anthropometric predictors (height (cm) and weight (kg)) and sex and age; 4) Model 4: Model 3 plus additional interaction terms between anthropometric variables and race/ethnicity; 5) Model 5: Model 4 plus all pairwise interactions between anthropometric variables, age, time between cancer diagnosis and LBM measurement, sex, and race/ethinicity.

Sex specific models were generated as follows: 1) Model 1: height (cm) and weight (kg) only; 2) Model 2: Model 1 plus age, time between cancer diagnosis and LBM measurement date, and race/ethnicity; 3) Model 3: Model 2 plus interaction terms between anthropometric predictors (height (cm) and weight (kg)) and age; 4) Model 4: Model 3 plus additional interaction terms between anthropometric variables and race/ethnicity; 5) Model 5: Model 4 plus all pairwise interactions between anthropometric variables, age, time between cancer diagnosis and LBM measurement, sex, and race/ethinicity.

To generate sex-by-race/ethnicitiy-specific predictive models, we constructed four different models among sex-by-race/ethnicitiy subgroups: 1) Model 1: height (cm) and weight (kg) only; 2) Model 2: Model 1 plus age, time between cancer diagnosis and LBM measurement date; 3) Model 3: Model 2 plus interaction terms between anthropometric predictors (height (cm) and weight (kg)) and age; 4) Model 5: Model 3 plus all pairwise interactions between anthropometric variables, age, time between cancer diagnosis and LBM measurement. Beyond height and weight, we also included other anthropometric measures in the models specified above in sensitivity analysis to test whether the inclusion of these other anthropometric variables would yield higher prediction accuracy.

#### Training and testing of algorithms

Data were split randomly into 75%: 25% for training and testing, respectively. For each imputed dataset, a series of linear regression models with least absolute shrinkage and selection operator (LASSO) penalty for variable selection were fitted, with a bootstrap procedure to obtain confidence intervals and p-values^40^. The LASSO was used to develop accurate but parsimonious predictive equations^40^. Results were aggregated across the twenty-five imputed datasets to obtain final coefficient estimates. The combination rule defined by Schomaker et. Al. (2017) was used to combine the bootstrap results across all 25 imputed datasets and construct (95%) confidence intervals (CIs) that take into account both the variability of variable selection and imputation^41^. To measure model fit, root mean square errors (RMSE) and correlation coefficients (R^2^) were calculated on the testing data for the overall sample as well as by sex and race/ethnicity subgroups. The entire training and testing procedure was repeated 10 times and the test set RMSE and R^2^ was averaged across all 10 replications for each model.

#### Correlations between predicted biomarkers of muscle mass and LBM

We tested and compared the strengths of correlations (R^2^) between urine creatinine levels and predicted LBM vs that between urine creatinine levels and DXA-based LBM.

## RESULTS

### Study Sample Population Characteristics

The distribution of participant characteristics in the overall study population and by sex are presented in **Table 1**. We identified 1777 participants with a self-reported cancer diagnosis with a median age of 71 (IQR:60 – 80). There were 933 (52.5%) females and 844 (47.5%) males. About 23.2% of the population were of a racial/ethnic minority background: Black (n=218, 12.3%), Mexican American and Hispanic (n=162, 9.1%), and Other (n=33, 1.9%). Men (55.9kg, SD=9.5) had higher DXA-based LBM than women (41.1kg, SD=7.4). Men were heavier (83.8kg vs. 73.1kg) and taller than women (174.3cm vs. 160.7cm).

**Table 1.**
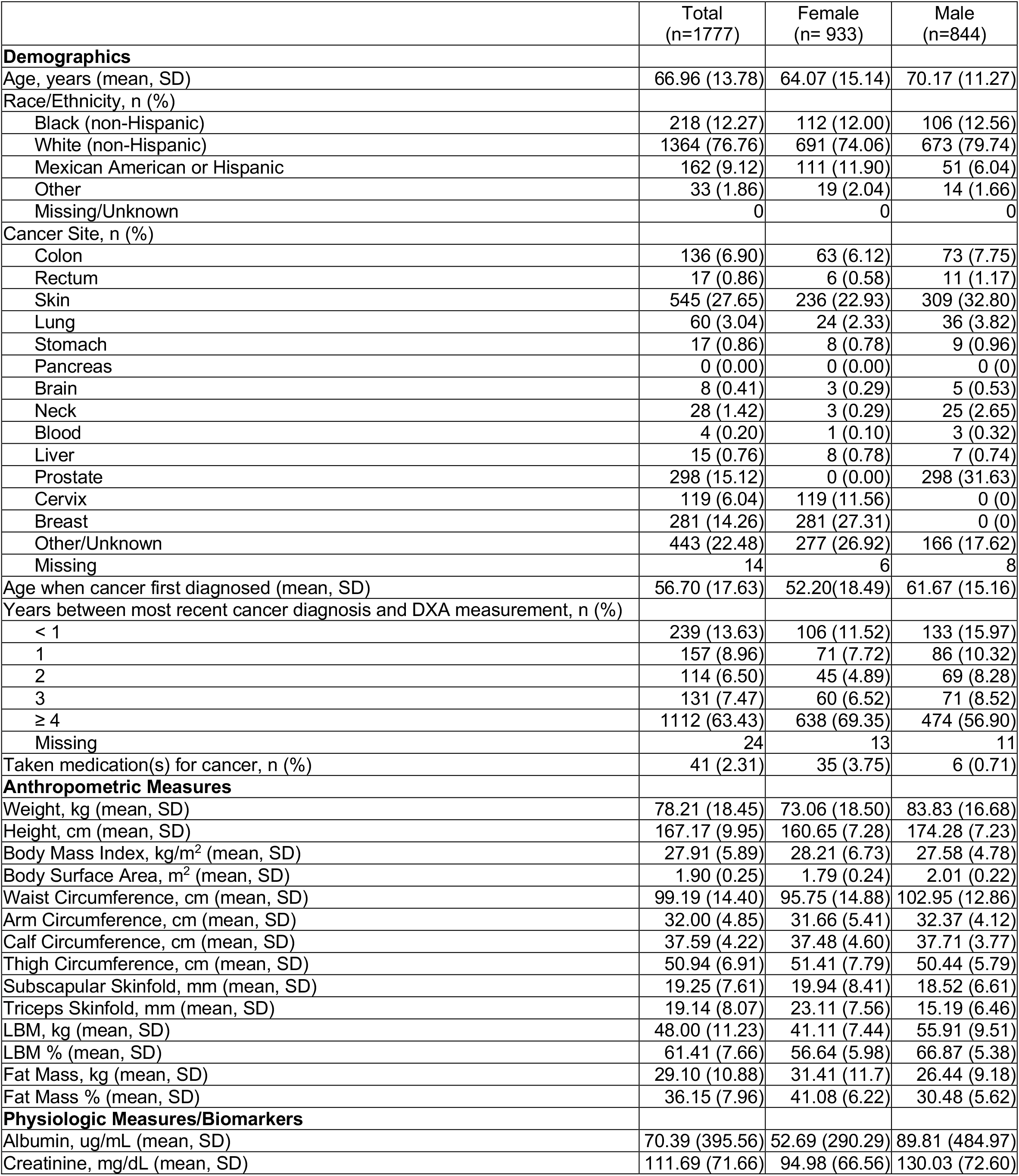
Distribution of baseline characteristics between men and women with at least one self-reported prior diagnosis of cancer, NHANES Survey Cycles 1999-2006

### Algorithm Performance

**Table 2** summarizes the performance of the primary model series overall, by sex, and sex by race stratified models based on R^2^ and RMSE values. In the overall sample, Model 1 was considered the most parsimonious model with high predictive accuracy (R^2^=0.86 and RMSE=4.26). Also, Model 1 had the best prediction accuracy with the fewest number of predictors (2) among females, (R^2^=0.93 and RMSE=2.46) and males, (R^2^=0.94 and RMSE=3.39) and among sex-by-race/ethnicity subgroups.

**Table 2.** Measures of model fit for primary anthropometric prediction equations for total LBM.

|  | **Model 1\*** |  | **Model 2<sup>§</sup>** |  | **Model 3<sup>+</sup>** |  | **Model 4<sup>†</sup>** |  | **Model 5<sup>‡</sup>** |  |
| --- | --- | --- | --- | --- | --- | --- | --- | --- | --- | --- |
|  | RMSE<sup>++</sup> | R<sup>2††</sup> | RMSE<sup>++</sup> | R<sup>2††</sup> | RMSE<sup>++</sup> | R<sup>2††</sup> | RMSE<sup>++</sup> | R<sup>2††</sup> | RMSE<sup>++</sup> | R<sup>2††</sup> |
| All | 4.26 | 0.86 | 2.99 | 0.93 | 2.81 | 0.94 | 2.81 | 0.94 | 2.86 | 0.94 |
| All Females | 2.64 | 0.93 | 2.59 | 0.93 | 2.61 | 0.93 | 2.61 | 0.93 | 2.61 | 0.93 |
| Black | 3.78 | 0.77 | 3.84 | 0.76 | 4.31 | 0.69 |  |  | 3.85 | 0.76 |
| White | 2.54 | 0.93 | 2.49 | 0.93 | 2.55 | 0.93 |  |  | 2.52 | 0.93 |
| Mexican-American or Hispanic | 2.26 | 0.95 | 2.57 | 0.93 | 2.90 | 0.92 |  |  | 2.56 | 0.93 |
| Other | 3.66 | 0.83 | 3.95 | 0.81 | 4.96 | 0.67 |  |  | 3.81 | 0.83 |
| All Males | 3.39 | 0.94 | 3.13 | 0.95 | 3.20 | 0.95 | 3.20 | 0.95 | 3.23 | 0.95 |
| Black | 3.59 | 0.94 | 3.21 | 0.95 | 3.81 | 0.93 |  |  | 3.2 | 0.95 |
| White | 3.44 | 0.94 | 3.17 | 0.95 | 3.21 | 0.95 |  |  | 3.24 | 0.95 |
| Mexican-American or Hispanic | 3.72 | 0.88 | 3.63 | 0.89 | 4.61 | 0.82 |  |  | 3.45 | 0.90 |
| Other | 3.95 | 0.83 | 4.43 | 0.79 | 8.58 | 0.24 |  |  | 4.00 | 0.80 |

\* Model 1 predictors are height (cm) and weight (kg) only.
§ Model 2 predictors are height (cm), weight (kg) and age.
+ Model 3 predictors are Model 2 predictors plus time interval between cancer diagnosis and DXA scan, sex, race/ethnicity.
† Model 4 predictors consist of Model 3 predictors plus additional interaction terms between anthropometric variables and race/ethnicity.
‡ Model 5 predictors are Model 4 predictors plus all pairwise interactions between anthropometric variables, age, time interval between cancer diagnosis and DXA scan, sex, and race/ethnicity.
++ RMSE = Root Mean Square Error
†† R<sup>2</sup> = R-squared

### Algorithms

**Table 3** shows the regression coefficients, of the most parsimonious model to predict DXA-based LBM (kg), overall, by sex and sex-by-race/ethnicity subgroups. Predictors for the overall model consist of weight (kg) (*β*=0.36; p-value<0.001) and height (cm) (*β*=0.52 p-value<0.05). For females, predictors for the most parsimonious model are weight (kg) (*β*=0.32; p-value<0.001) and height (cm) (*β*=0.17; p-value=0.15). For males, predictors for the most parsimonious model are weight (kg) (*β*=0.41; p-value<0.001), height (cm) (*β*=0.32; p-value=0.12). The most parsimonious model for Black, non-Hispanic females, White, non-Hispanic females, Mexican-American or Hispanic females, other females, Black, non-Hispanic males, White, non-Hispanic males, Mexican-American or Hispanic males and Other, non-Hispanic males included the same predictors. Similar results were observed when additional anthropometric variables were included to build a series of secondary algorithms.

**Table 3.** Regression coefficients for the most parsimonious models overall, by sex, and by sex and race/ethnicity of the associations between anthropometric measures and total LBM.

| Categories | Algorithms |
| --- | --- |
| All | $LBM = -67.74 + 0.36^{\ddagger} * (Weight) + 0.52^{\dagger} * (Height)$ |
| All Females | $LBM = -10.64 + 0.32^{\ddagger} * (Weight) + 0.17 * (Height)$ |
| Black | $LBM = 1.23 + 0.31 * (Weight) + 0.12 * (Height)$ |
| White | $LBM = -9.37 + 0.32^{\ddagger} * (Weight) + 0.17 * (Height)$ |
| Mexican-American or Hispanic | $LBM = -13.25 + 0.34^{\S} * (Weight) + 0.18 * (Height)$ |
| Other | $LBM = -37.29 + 0.35^{\dagger} * (Weight) + 0.33 * (Height)$ |
| All Males | $LBM = -33.46 + 0.41^{\ddagger} * (Weight) + 0.32 * (Height)$ |
| Black | $LBM = -31.05 + 0.46^{\ddagger} * (Weight) + 0.29 * (Height)$ |
| White | $LBM = -33.45 + 0.41^{\ddagger} * (Weight) + 0.32 * (Height)$ |
| Mexican-American or Hispanic | $LBM = -34.38 + 0.43^{\dagger} * (Weight) + 0.32 * (Height)$ |
| Other | $LBM = -58.82 + 0.36^{\dagger} * (Weight) + 0.49 * (Height)$ |

‡ P-value $\leq 0.001$
$\S$ P-value $\leq 0.01$
† P-value $\leq 0.05$

The correlations between urine creatinine levels and LBMs (DXA-based and predicted) are visually represented in **Figure 2**. The magnitude of correlations between derived LBM-algorithm and urine creatinine levels were large compared to those measured between DXA-based LBM and urine creatinine levels (R^2^=0.30 vs. R^2^=0.17) The correlations between predicted LBM measures with urine creatinine levels were comparable to those observed for DXA-based LBM among females (R^2^=0.25 vs R^2^=0.23) and among males (R^2^=0.17 vs R^2^=0.17). These correlations were similar among sex-by-race/ethnicity subgroups. Therefore, validating our conclusions that the sex by race/ethnicity stratified Model 1 derived algorithm of LBM (kg) is a sufficient measure for estimating DXA measured LBM (kg).

## DISCUSSION

In this large population-based study, we developed and validated predictive algorithms comprised of simple and routinely measured anthropometric measurements among cancer patients. Sex, race/ethnicity, and time between cancer diagnosis and DXA measurement were also added as predictors to improve accuracy among these subgroups of cancer patients. To the best of our knowledge, there are no previously published data on the derivation of algorithms for predicting LBM among cancer patients using routinely measured anthropometric measurements. The developed algorithms will provide clinicians with a tool to estimate the total LBM composition of cancer patients prior to initiation of chemotherapy based on routinely measured anthropometric measures which are non-invasive and inexpensive.

In the non-cancer population, several algorithms to predict body compositions including LBM have been developed and validated^23, 24^. Lee et. al. (2017) recently developed predictive equations using data from NHANES participants (with or without cancer diagnosis)^23^. In this study, a statistical model including age, height, weight, waist circumference, arm circumference, calf circumference, thigh circumference, triceps skinfold, subscapular skinfold, race, accurately predicted LBM in men (R^2^=0.94) and women (R^2^=0.87)^23^. In comparison, our most parsimonious model accurately predicted LBM of cancer patients with the same number of features and to a similar degree of fit as Lee et al (2017)^23^. Finally, the predictors of the models were limited to the anthropometric measures collected in NHANES. Published literature supports the use of tricep and subscapular skinfold in conjunction with circumference measures to accurately measure body composition ^42^.

## Conclusion

Prediction equations utilizing anthropometric predictors of height and weight to estimate DXA-based LBM composition of cancer patients were developed, tested and validated. Models were further improved with sex, race/ethnicity, and time interval between cancer diagnosis and LBM measurement as predictors. These equations will simplify the ability of clinicians to estimate the DXA-based LBM composition of cancer patients prior to initiation of chemotherapy based on routinely measured anthropometric measures which are non-invasive and low cost to patients. Our next step is to apply these derived equations to measure LBM thresholds associated with the risk of chemotherapy adverse events.

## Data Availability

All data referred to in the manuscript is publicly available via CDC.

https://www.n.cdc.gov/nchs/nhanes/Default.aspx

